# Streptococcus pneumoniae: Nasal influenza vaccination, carriage density and transmission in families

**DOI:** 10.1101/2024.04.25.24306231

**Authors:** J. Metz, G. Qian, B. Morales-Aza, J. Oliver, E. Oliver, H. Rice, K. Duale, P. Heath, S.N. Faust, M.D. Snape, S. Hughes, L. Hole, R. Mann, F. Shackley, P. Rudd, S. Ludman, BD. Gessner, L. Danon, A. Finn

## Abstract

The live attenuated influenza vaccine (LAIV) is offered in the United Kingdom to young children, protecting against influenza for those vaccinated and indirect protection for the wider community. It has also been shown to increase carriage density of Streptococcus pneumoniae, to an extent, in children. This study therefore investigates whether the vaccine leads to an increase in density in children and, if so, whether this augments transmission to household contacts.

We implemented a randomised control study involving 405 two-year-old children and 958 household contacts. Nasopharyngeal swabs from all participants were taken over 5 visits, each two weeks apart, and tested for pneumococcal carriage. LAIV was given to 205 children at visit 1, and to 200 children at visit 2.

We developed regression models to analyse the association between vaccination and whether an increase in pneumococcal density 14 and 28 days later was observed, as well as an increase in the odds of transmission to household members after administering LAIV.

From regression analyses, there was a 2.5-fold (95%CI:1.3-4.3, p<0.001) increase in the odds of vaccinated children to have increased pneumococcal density 2 weeks later, compared with unvaccinated children, and a 2.1-fold (95%CI:1.2-3.6, p=0.011) increase in the odds of presumed transmission from children to their household contacts.

Our results provide evidence that an attenuated influenza virus infection transiently increases the likelihood of pneumococcal transmission from children who are colonised with the bacterium to their contacts and that this increase is driven by an increase in bacterial abundance triggered by the vaccine.

## Introduction

Despite the global disease burden of *Streptococcus pneumoniae* (Sp), factors determining Sp transmission have not been fully evaluated. Substantial evidence supports that upper respiratory tract (URT) pneumococcal colonisation sustains population-based transmission^1,2^ and Sp transmission requires close proximity.^1^ Young children are most likely the main reservoir and transmission drivers of Sp: they carry Sp at highest rates, particularly if attending daycare; carers of young children carry Sp at higher rates than non-carer adults of similar age; and high vaccination coverage against Sp in early childhood with pneumococcal conjugate vaccines (PCV) reduces vaccine-type disease and carriage in the whole population.^3,4^

There have been few studies evaluating household Sp transmission and, to our knowledge, none investigating the impact of Sp density on household transmission. The intra-nasal LAIV is offered every autumn in the UK childhood immunisation programme to children aged 2 years and over. LAIV provides effective protection against influenza for those vaccinated and indirect protection for the wider community.^5^ LAIV has also been shown to increase Sp carriage density in mice and, to a limited degree, in children.^6–8^ This attenuated upper respiratory tract (URT) infection can therefore be used to investigate whether increases in Sp density in carriers augment transmission to their contacts. We conducted a randomised controlled trial to assess the impact of LAIV administration on Sp carriage density in previously LAIV-naive 2-year-old children and onward transmission to their household contacts.

## Methods

In this multi-centre prospective randomised control trial conducted in the UK, all samples were collected between September and December 2017 and October and December 2018.

### Ethical approval

Ethical and governance approvals were granted by the UK Research Ethics Committee (Cambridgeshire and Hertfordshire) (17/EE/0351) and Health Research Authority, as well as local NHS Research and Development Departments at each site. All participants or their parents/carers provided written informed consent either in writing or by an electronic process.

### Funding and sponsorship

The study was sponsored by the University of Bristol and jointly funded by the Bill & Melinda Gates Foundation and Pfizer.

### Study design

#### Recruitment and enrolment

43544 letters inviting study participation were sent to eligible households within the vicinity of the 10 study sites via the local child health records offices that organise routine childhood vaccination appointments. The study sites were Bristol, Oxford, London, Southampton, Manchester, Taunton, Gloucester, Sheffield, Bath and Exeter. 405 household family units were fully consented and enrolled into the study early in the LAIV immunisation period (September-October) and the study was carried out from October to December during both 2017 and 2018. An eligible household family unit was defined as a 2-year-old index child due their first LAIV dose and at least two household contacts of any age.

### Randomisation, intervention and procedures

We implemented a randomised control study with a stepped-wedge-like design. All participating households had five study visits (visits 1 to 5) either at home or in clinic at 14-day intervals to collect NP samples from each participant at each visit (Figure S1). Demographic data as reported by parents including age, gender, household size, day-care/school attendance and vaccination status of the index child were collected at baseline and clinical data on current/recent antibiotic use and self-assessed coryzal status according to the ‘Symptoms of Nasal Outflow Tally’ (scored 0-3) as previously defined^9^ were collected at each study visit. At enrolment (day 0, visit 1), household family units were randomised 1:1 into either the early LAIV group, in which the 2-year-old index child was immunised with LAIV at visit 1 (after baseline sample collection), or to the control late LAIV group where the index child did not receive LAIV until 28 days later, at visit 3 (after visit 3 sample collection). Household members eligible for LAIV, other than the index child, were asked to defer LAIV until all five visits and NP samples had been collected. Three families withdrew prior to completion of visit 1 and therefore were not included in subsequent study analysis. 391 households (97%) made up of 1317 participants (96%), completed the study. The study was conducted during two consecutive LAIV-vaccinating seasons; 1985 samples from 422 participants were collected between October and December 2017 (season 1) and 4495 samples from 957 participants were collected between October and December 2018 (season 2). A summary of the numbers of families and participants enrolled, sampled at each visit, vaccinated, and analysed is shown in the consort flow diagram (Figure 1).

**Figure 1:**
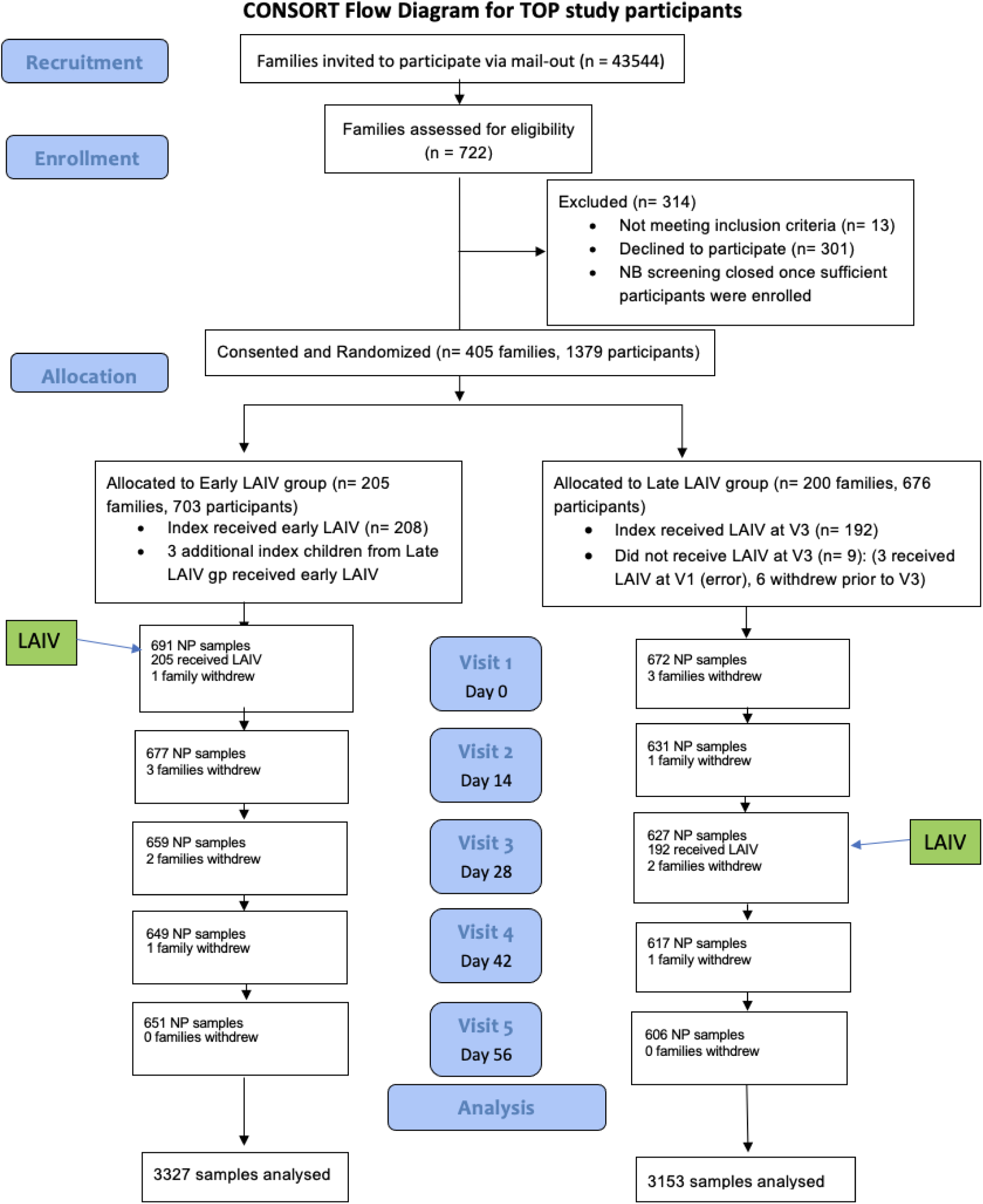
Consort Diagram. Of the 43,544 families invited to participate, 722 families were assessed for eligibility during the study period. 405 families were subsequently allocated to one of two study arms: the Early LAIV group, in which index children (205/205) received live attenuated influenza vaccine at visit 1, or the Late LAIV group, in which most index children (192/200) received the vaccine at visit 3. LAIV, live attenuated influenza vaccine; NP, nasopharyngeal.

### Laboratory Analysis

NP samples were taken with PurFlock Ultra swabs (Medical Wire & Equipment) using a standardised method (as detailed in the study protocol and standard operating procedure, supplement S1), transported in 1.5ml of STGG (at 2-8℃) and frozen within 4 hours. All samples were analysed for the presence of Sp by qPCR with lytA as the gene target as previously described with a cycle threshold (Ct) cut off of 35 and Ct values converted into gene copies per ml (gc/ml) using a standard curve.^7^ In this paper, lytA detection Ct<35 is referred to as Sp positivity.

### Statistical Analysis

Baseline comparisons between participant groups were carried out using all samples taken. An intention-to-treat (ITT) analysis utilising all available data from participants with valid consent, irrespective of whether participants provided a sample at every visit, was used for Sp point prevalence and index density plotting. Results from children and their contacts with complete data (results available from every visit) were used for the per-protocol analysis and index children positive for Sp at baseline together with their discordant (negative for Sp at baseline) contacts were included for parts of the transmission analysis. Prevalence proportions are reported as percentages with associated binomial confidence intervals (CIs). The Mann-Whitney and Kolmogorov-Smirnoff tests were used to assess whether the change in Sp density between visits 1 and 2 in index children who were in the early LAIV group was statistically different from changes in density in index children in the control late LAIV group.

We employed two complementary approaches to analyse the data. First, we used statistical tests. Chi-squared test was used to compare Sp carriage point prevalence and transmission data, and Mann-Whitney-U test was used to compare densities. To ensure robustness of results, and take into account the wide natural variability in carriage density,^7^ we also performed regression analyses: we developed a simple logistic regression model to analyse the association between vaccination with LAIV (binary explanatory variable) and whether an increase in pneumococcal density 14 and 28 days later was observed (categorical outcome variable). A Chi-squared test was used to assess whether there was a change in the odds of transmission to household members after administering LAIV. This was subsequently confirmed using a logistic regression model. Finally, mixed-effects models of both density and transmission, with random intercepts to account for the random effects among index children, were developed as a sensitivity analysis. All analyses were carried out in R version 4.2.3.

## Results

### Demographics and pneumococcal carriage at baseline

Of the 1379 enrolled participants, 1363 participants provided a baseline (visit 1) sample, including 550 children aged less than 5 years (of which 405 were index participants), 159 children aged 5 to 17 years and 654 adults.

Visit 1 occurred between 3^rd^ October – 31^st^ October 2017 or 27^th^ September – 15^th^ November 2018. Young children more frequently carried Sp and at higher densities than older household members (Figure 2). 276/405 (68%) index children (2 years of age) and 227/958 (24%) contacts were carriers at baseline. Antibiotic use (current or within the past week) was minimal across all participants (Table 1a and 1b).

**Figure 2:**
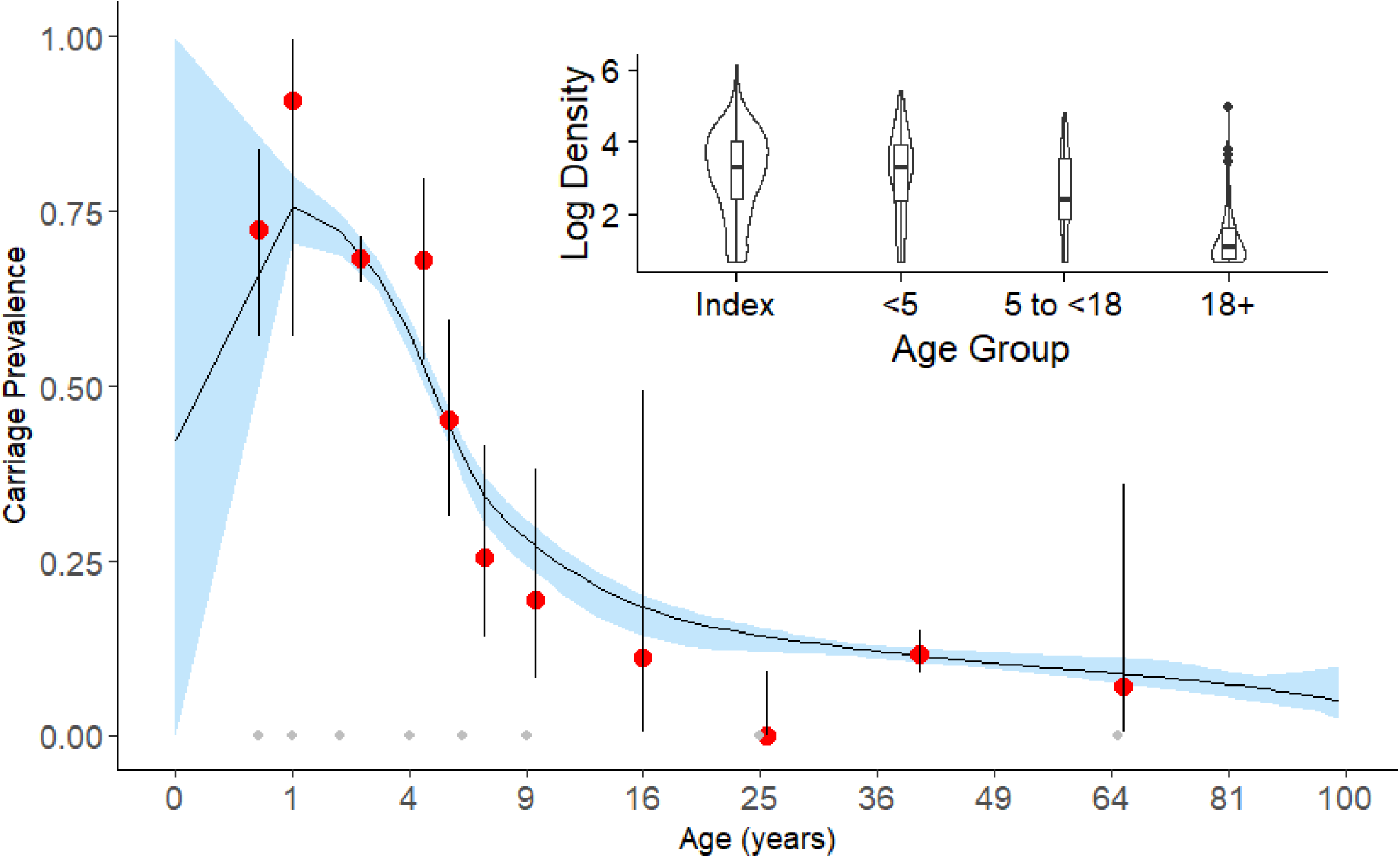
Pneumococcal carriage and density (inset) as a function of age. Red dots represent the observed carriage point prevalence, black vertical bars the 95% binomial confidence intervals, black curve the spline fit and blue shaded area the 95% credible intervals of the spline. Inset: Violin plots show the log carriage density among index children and their contacts at visit 1. The boxplots nested within show the quartiles of these log densities. Methodology for the spline fit and Bayesian fitting can be found elsewhere.^12^

**Table 1a:** Comparing demographic/clinical characteristics between index participants at baseline. Note, 3 index participants are not included in this table as they were missing demographic/clinical information. Statistical analysis testing for categorical data(*) was done using the chi-square test and for continuous data the independent t-test if normally distributed (∼) and the Mann-Whitney test for non-normally distributed data (§). LAIV, Live Attenuated Influenza Vaccine; SNOT, Symptoms of Nasal Outflow Tally, where 0 = no coryzal symptoms, 1 = mild, 2 = moderate and 3 = severe; Abx Score, antibiotic score where 1 = no abx, 2= abx in the past 7 days and 3= current abx.

| Characteristic | Group | Early LAIV |  | Control Late LAIV |  | P-value |
| --- | --- | --- | --- | --- | --- | --- |
|  |  | N | Value | N | Value |  |
|  |  | 202 (205) |  | 200 |  |  |
| Age in years | Median | 2.47 | 2.47 | 2.50 |  | 0.82 |
| Gender) | Male | 86 | 42% | 108 | 54% | 0.02** |
|  | Female | 116 | 58% | 91 | 46% |  |
| Daycare attendance | Count | 150 | 74% | 150 | 75% | 0.8 |
| Household size § | Mean | 3.94 |  | 3.92 |  | 0.84 |
| SNOT score§ | Mean | 0.619 |  | 0.670 |  | 0.71 |
| SNOTTY* (SNOT score > 0) | Positive | 102 | 51% | 99 | 50% | 0.9 |
| Abx score § | Mean | 1.02 |  | 1.04 |  | 0.51 |

**Table 1b:** Comparing demographic/clinical characteristics between contact participants at baseline. Note, 3 index participants are not included in this table as they were missing demographic/clinical information. Statistical analysis testing for categorical data(*) was done using the chi-square test and for continuous data the independent t-test if normally distributed (∼) and the Mann-Whitney test for non-normally distributed data (§). LAIV, Live Attenuated Influenza Vaccine; SNOT, Symptoms of Nasal Outflow Tally, where 0 = no coryzal symptoms, 1 = mild, 2 = moderate and 3 = severe; Abx Score, antibiotic score where 1 = no abx, 2= abx in the past 7 days and 3= current abx.

| Characteristic | Early LAIV<br><5 yo<br>contacts | Late LAIV<br><5 yo<br>contacts | P- value | Early LAIV<br>5-17 yo<br>contacts | Late LAIV<br>5-17 yo<br>contacts | P- value | Early LAIV<br>>18 yo<br>contacts | Late LAIV<br>>18 yo<br>contacts | P- value |
| --- | --- | --- | --- | --- | --- | --- | --- | --- | --- |
| N | 73 | 69 |  | 76 | 74 |  | 322 | 309 |  |
| Age (years) ~<br>(median) | 2.6 | 3.8 | 0.4 | 6.7 | 6.5 | 0.4 | 37.4 | 36.5 | 0.06 |
| Gender*<br>M:F (% male) | 41:32<br>(56%) | 30:38^<br>(44%) | 0.2 | 36:39^<br>(48%) | 34:39^<br>(47%) | 0.9 | 117:205<br>(36%) | 113:195<br>^<br>(37%) | 0.9 |
| Daycare<br>* N (%) | 13 (18%) | 13 (19%) | 0.9 | 0 | 0 | / | 1 (0.3%) | 4 (1.3%) | 0.2 |
| Household<br>size §<br>(mean) | 4.3 | 4.22 | 0.6 | 4.53 | 4.46 | 0.8 | 3.91 | 3.9 | 0.9 |
| SNOT<br>score §<br>(mean) | 0.53 | 0.67 | 0.4 | 0.4 | 0.36 | 0.6 | 0.26 | 0.35 | 0.2 |
| SNOTTY<br>*<br>i.e.<br>SNOT<br>score > 0<br>(%) | 31<br>(43 %) | 32<br>(46%) | 0.6 | 26<br>(34%) | 21<br>(28 %) | 0.5 | 75<br>(23%) | 85<br>(28%) | 0.2 |
| Abx<br>score §<br>(mean) | 1.07 | 1.03 | 0.7 | 1.03 | 1 | 0.2 | 1.03 | 1.04 | 0.2 |

Sp density followed a skewed distribution in index children, with variation over six orders of magnitude: 3.5 x 10^0^ to 1.3 x 10^6^gc/ml with an interquartile range of 10,137gc/ml (288 - 10424gc/ml). Gene copies per ml were plotted on a log_10_ scale and medians were used for comparisons (Figure 2).

Comparing study arms at baseline (prior to any intervention), more than two thirds of index children were Sp carriers with identical prevalences in those assigned to early or late LAIV: 139/205 (68%, 95% CI 61-74%) in the early LAIV group and 136/200, (68% 95% CI 61-74%) in the control late LAIV group were carriers. Just under a quarter of household contacts were Sp carriers, across both study groups with similar rates: 100/486 (23% 95% CI 19-27%) in the early LAIV group and 118/472 (25% 95% CI 21-29%) in the control late LAIV group were carriers.

At baseline, contacts of Sp positive index children were significantly more likely also to be Sp carriers than contacts of Sp negative index children, 181/652 (28%) compared with 46/306 (15%) respectively, Chi-square test, p-value = 0.0001. The relative risk of the contact being positive for Sp if the index child was positive was 1.85 (95% CI 1.38 to 2.49, Chi-square test).

### Randomisation and elimination of potential confounders

There were no statistically significant differences in demographics between the two study groups neither among index nor contact participants, apart from index child sex (control late LAIV group 54% males, early LAIV group 42%) (Tables 1a & 1b). Since sex was not associated with Sp carriage point prevalence nor density, no additional statistical adjustments were made.

### Impact of LAIV on pneumococcal carriage in recipients

An increase in density was observed in 62/128 (48%, 95%CI: 40-57%) of the index children who had received LAIV at visit 1, 14 days post-immunisation, and in 32/117 (27%, 95%CI: 20-36%) of those who had not received LAIV (Chi-square test p=0.0007) (Figure 3). This was confirmed by both fixed and mixed-effects logistic regression (Tables 2 and S1). Augmenting the fixed-effects simple logistic regression model to include random effects among different index children showed a similar odds ratio to that seen with the fixed effects model; the Akaike Information Criterion (AICs) for the fixed and mixed-effects models were 319 and 321, respectively. In addition, the magnitude of these density increases tended to be larger in the group that had received LAIV than in the group that had not (Figure 4). Including only index participants that were Sp carriers at both visit 1 and 2, the median change in density between visit 1 and 14 days later at visit 2 was +854gc/ml in LAIV recipients (early LAIV group) and -393gc/ml in LAIV non-recipients (control late LAIV group), p = 0.04 by Mann Whitney U test and p = 0.005 by Kolmogorov-Smirnov test. By visit 3, 28 days following LAIV, there was little evidence of a persistent increase in Sp density relative to baseline (visit 1) among LAIV recipients when compared to non-recipients (Tables 2 and S1). These data indicate that LAIV achieved the intended outcome of transiently increasing Sp density, allowing us to further assess the impact of LAIV in the context of this increase on household transmission.

**Figure 3:**
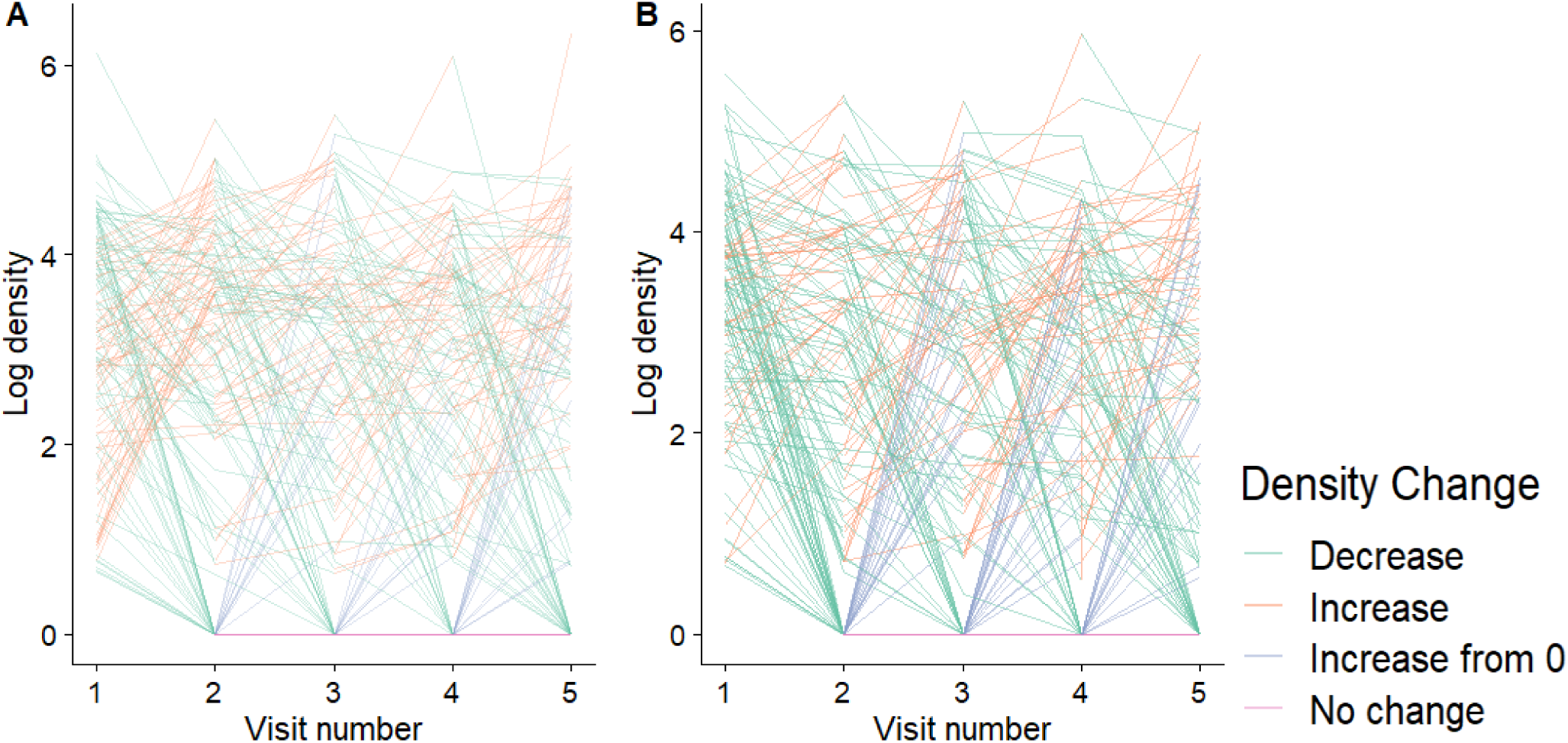
The dynamics of pneumococcal carriage density in visit 1-positive index children across all 5 visits in A. early LAIV group (n = 128) and B. control late LAIV group (n = 117). Green lines denote a decrease in density, blue lines denote an acquisition event (increase from 0) and red lines denote an increase. An increase in density was observed in 62/128 (48% 95%CI 40-57%) of the index children who had received LAIV, at visit 2 14 days later, and in 32/117 (27% 95%CI 20-36%) of those who had not received LAIV (Chi-square test p = 0.0007) (Figure 3). This was confirmed by both fixed and mixed-effects logistic regression (Tables 2 and S1): augmenting the fixed-effects simple logistic regression model to include random effects among different index children showed a similar odds ratio observed from the fixed effects model; the AIC for the fixed and mixed-effects models were 345 and 347, respectively. Note: this was a per-protocol analysis including participants that had provided a sample at each study visit.

**Figure 4:**
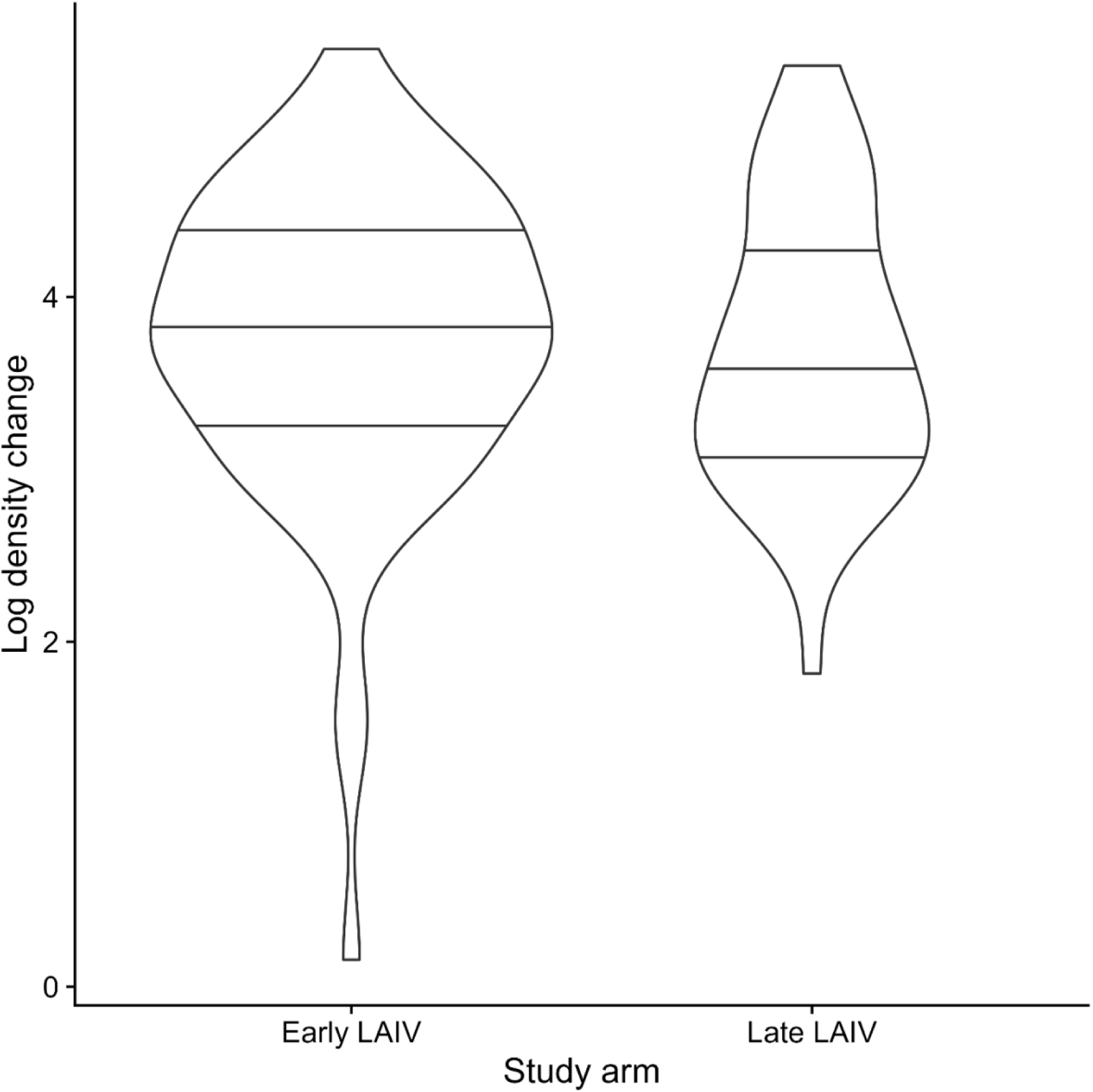
Change in pneumococcal carriage density by arm from visit 1 to visit 2, with index children from the early LAIV group on the right and control late LAIV group on the left. The median and interquartile ranges are shown width of the violin plots inside each violin. The numbers of children who carried pneumococcus at visit 1 and visit 2 and whose carriage density increased between these 2 visits in each group are represented by the width of the violin plots.

**Table 2:**
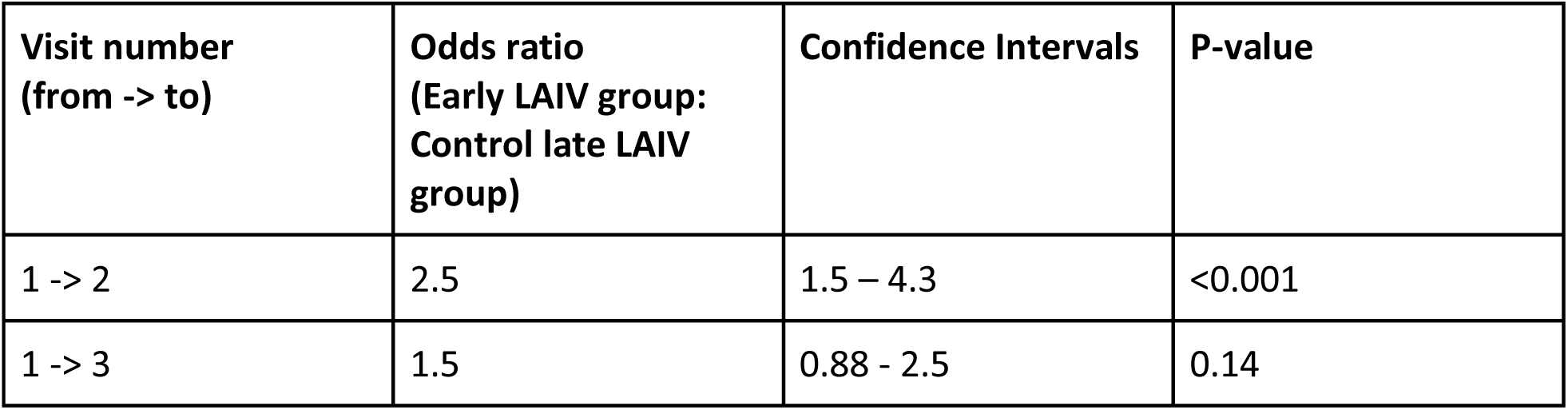
Odds ratio between index children in the early LAIV group and control late LAIV group of an increase in density in these children, from visit 1 to visit 2, and visit 1 to visit 3, by univariate logistic regression.

### Impact of LAIV on transmission

Considering all baseline pneumococcal carriage-negative contacts, irrespective of the index child’s baseline carriage status, in the early LAIV group, 59/367 (16%, 95% CI 13-20%) became positive by 14 days after LAIV vaccination of the index child. In the control late LAIV group, 31/329 (9%, 95% CI 7-13%) contacts became positive within the same time period, (chi-squared test, p = 0.006). By the time of the third study visit at 28 days (when Sp carriage density was similar among index children that had and had not received LAIV) there was no discernible difference between contact Sp acquisition rates in the two study groups: 16% (early) and 15% (late), p = 0.7).

When we restricted analysis to baseline Sp carriage positive index children and their contacts who were baseline Sp carriage negative, 63/437 (15%) of the contacts were Sp positive at visit 2, 14 days later including 45/237 (19%) in the early LAIV group and 21/205 (10%) in the late LAIV group, p= 0.015. Chi-squared and both fixed and mixed-effects regression analyses of transmission in these families showed that the odds of detecting Sp transmission from index children to household members was around 2 times greater 14 days after they had received LAIV than among control families (Tables 3 and S2). Augmenting the simple logistic regression model to include random effects among different discordant pairs showed a similar odds ratio to that seen with the fixed effects model; the AIC of the former was 372, while that of the latter was 370.

**Table 3:** Univariate logistic regression analysis comparing transmission events from 2-year-old index children carrying pneumococcus at baseline to family contacts at visit 2 (14 days) and visit 3 (28 days) between the two study groups. At visit 3, we considered family contacts only if they had been Sp-negative at both visits 1 and 2. Discordant pairs were defined as an index child who was an Sp carrier and household contact who was Sp-negative at a particular visit.

| Transmission events<br>between: | Odds ratio | Confidence Intervals | P-value |
| --- | --- | --- | --- |
| Discordant pairs at<br>visit 2 | 2.1 | 1.2 - 3.6 | 0.011 |
| Discordant pairs at<br>visit 3 | 0.89 | 0.53 - 1.5 | 0.67 |

We considered the possibility that LAIV increased transmission independently of causing increases in Sp carriage density. To evaluate this, we assessed the median carriage density of baseline-positive index participants at visit 2 whose contacts had become positive at visit 2 and compared these between the two study groups. We found that Sp density was similar between the two groups, with a median density of 4744gc/ml in the late LAIV group and 5657gc/ml in the early LAIV group, Mann Whitney U test, p=0.64. These data support the hypothesis that Sp density was the main driver of increased household transmission.

By visit 3, on day 28, there was no significant difference (p= 0.8) in Sp carriage point prevalence between contacts of LAIV recipients (early LAIV group) and non-recipients (control late LAIV group) (35/226, 15%, 95%CI: 11-21 and 34/200 17%, 95%CI: 12-22%, respectively). Both fixed and mixed-effects logistic regression showed that there was no significant difference between the rates of transmission events detected at visit 3 (28 days) between the two study groups among families whose index 2-year-old child carried pneumococcus at baseline but whose contacts were negative at both visits 1 and 2 (Tables 3 and S2). We also found that, of the 66 contacts among the discordant pairs who were Sp positive at visit 2, 27 remained positive at visit 3; again, no significant difference between the early and late LAIV groups was observed between these contacts (p = 0.13, Table S3).

### Impact of pneumococcal density on transmission

As additional support for the data from the randomised control trial, we assessed across both study arms whether Sp carriage density predicted household transmission using an observational retrospective analysis of visit 2 samples. This analysis showed that the carriage density in index children where acquisition of Sp in one or more contacts had occurred was significantly higher than in index children where all contacts remained Sp carriage negative: the median densities were 5657gc/ml, compared with 322gc/ml respectively, (p< 0.001 from the Mann Whitney U-test). Separating study arms, a difference in density remained in both groups: in the early LAIV group, the median density in index children where at least one contact had acquired Sp was 5657gc/ml, compared with 739gc/ml in those whose contacts remained Sp negative (p=0.002 from the Mann Whitney U-test). In the late LAIV group index children (where LAIV had not yet been received), the median density was 4744gc/ml in transmitting index participants compared with 180gc/ml in non-transmitting index participants (p = 0.02 from the Mann Whitney U test), (Figure 5).

**Figure 5:**
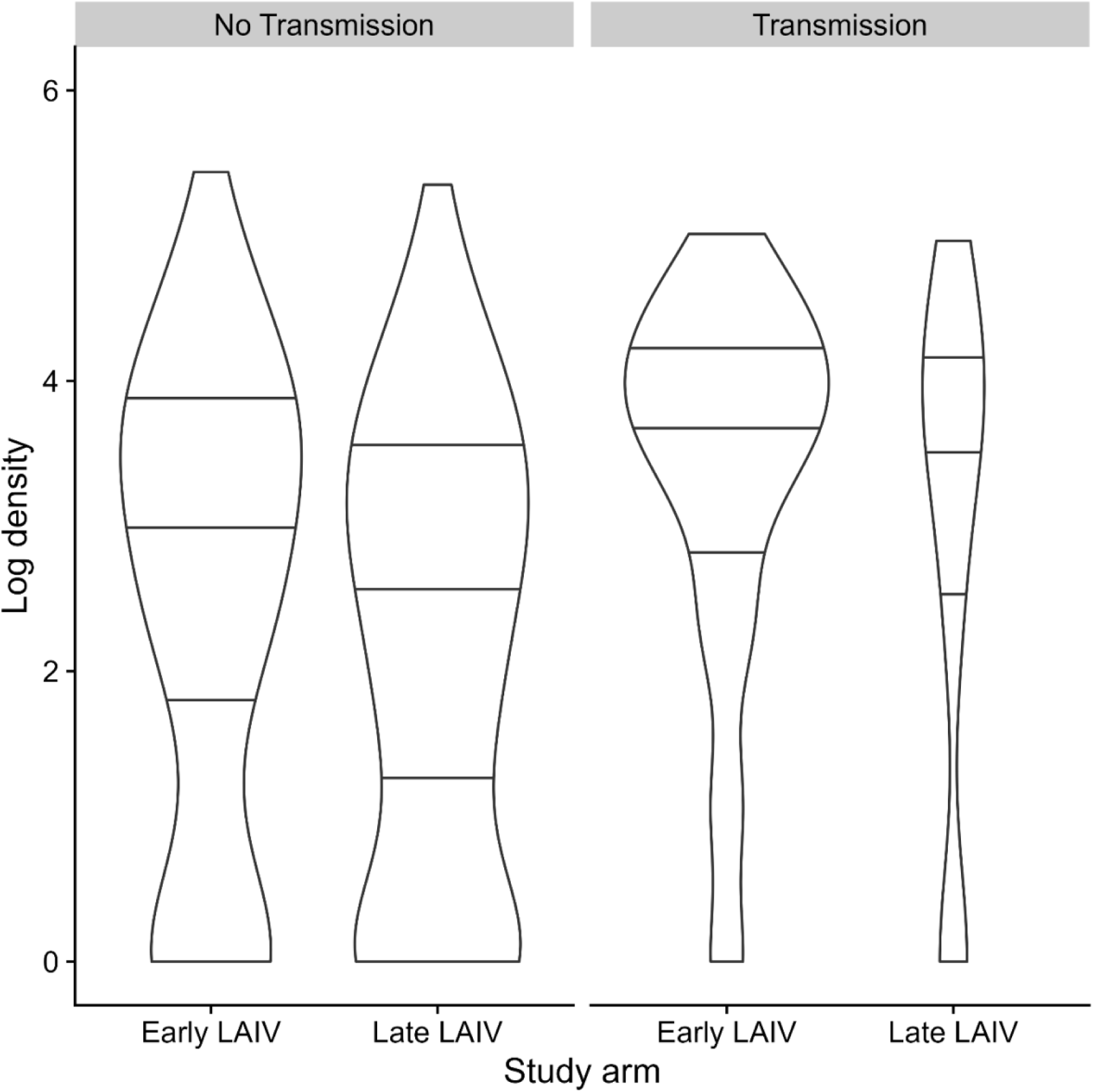
Carriage density of index children at visit 2 (14 days after LAIV in Early LAIV group) plotted according to whether a transmission event had occurred or not and separated by study group. Note that we used the transform: log (Density+1) on the y-axis. The width of the plots represents the number of events. There was a significant statistical difference, Mann Whitney U-test, p< 0.001, between the carriage densities of index children where there had been a transmission event, n = 62, median density 5657 gc/ml compared with where no transmission event had been captured, n = 185, median density 322 gc/ml. When this analysis was repeated but by study arm, a significant difference in pneumococcal densities was found in both groups: in the vaccinated early LAIV group, the median density of transmission-event positive index participants was 5657gc/ml (n = 44), compared with 739gc/ml (n = 87) in the transmission-event negative index, p = 0.002. In the unvaccinated late LAIV group; index pneumococcal carriage median density 4744gc/ml (n = 18) in transmission-event positive index compared with 180gc/ml (n = 98), in transmission-event negative index, p = 0.02.

## Discussion

As shown previously in studies from our group and others,^4,6,8^ intranasal administration of attenuated influenza virus strains transiently increased detectable bacterial load in individuals colonised with Sp. In our study we took samples at 14-day intervals and found this rise to occur at 14 days but to have largely dissipated by 28 days. This phenomenon raised the possibility that such inoculation with a combination of attenuated respiratory viruses might be used as a probe to explore whether such changes also influence transmission of nasal bacteria to close contacts of vaccine recipients, the primary aim of this experiment.

This was duly confirmed: we found that, in the context of a relatively modest rise in nasal Sp density, and when compared to controls who had not received LAIV, there were temporarily detectably higher odds of presumed Sp transmission from LAIV vaccinated children, as well as the subset who were Sp carriers, to their baseline Sp-negative contacts. This occurred notwithstanding the great background diversity of Sp carriage density that existed in young children. While it is plausible that the increased pneumococcal abundance rendered influenza vaccine recipients more infectious, it is also possible that other vaccine-induced changes, for example in bacterial phenotype or in host responses, contribute to enhancing the number and efficiency of transmission events.

Taken together, our results provide, for the first time, strong evidence that an attenuated influenza virus infection increases the likelihood of transmission of pneumococcus from young children who are colonised with the bacterium to their home contacts and that this increase is driven largely by an increase in bacterial abundance triggered by the influenza vaccine. This observation raises several new questions about the nature of the dynamics of the bacterial density - transmission relationship and the mechanisms involved, particularly how the bacteria sense the viral infection and whether the host response to the vaccine is involved. Some of our results, including the higher likelihood of Sp acquisition following receipt of LAIV and comparisons between the colonisation densities of transmitters and non-transmitters in the two study groups, suggest the possibility that the vaccine may also be having effects that influence transmission that are distinct from causing increased colonisation density. However, this conclusion is not clear cut from the evidence we present and will require further investigation.

The implications of these findings for pneumococcal ecology at the population level cannot be deduced from the results of this small study. On one hand, the observed vaccine-induced increases in colonisation density and rates of transmission were modest and transient as compared to the wide range of densities we observed and increases in bacterial densities that have been described in association with common wild-type respiratory viral infections.^10^ On the other hand, LAIV is administered to large numbers of children over a relatively short period of time within the last and first quarter of each year in the UK. Given the well-accepted importance of children in the dynamics of pneumococcal transmission across all age groups, this question deserves further attention.

The main strength of this study was in its randomised controlled design, eliminating potential biases and permitting causality to be inferred with regards to the findings. A limitation was detection of Sp only by its autolysin gene rather than also by serotype confirmation, making it less certain that a transmission event from study child to family member occurred for any individual child. At the full study population analytic level, the randomised design should have protected against bias consequent upon any such errors. The relative infrequency of sampling reduced the precision with which the density changes and transmission events could be tracked, and ideally more frequent testing would have occurred; however, we were constrained to a frequency that participating families would accept.

Transmission between discordant pairs from visit 1 to visit 2 was mostly limited to 1 event per household (Figure S3). Hence, a fixed effects model was found to be sufficient, with a lower AIC compared with the mixed effects model. However, the number of transmission events at visit 3 was much more widely distributed. Hence, the mixed effects model outperformed its fixed effects counterpart, in terms of their respective AIC values.

Aside from providing new insights into the complex interactions between viruses, bacteria and the human host in the URT, the findings of this study add credence to the hypothesis that the control of symptomatic and invasive infections caused by colonising pathobiont bacteria may be achieved to some degree through prevention of respiratory viral infections with mucosal vaccines, assuming vaccines can provide sterilizing immunity or, more likely, reduction of viral load during infection. Our results in turn are consistent with observations about the relationships between respiratory viral infections and bacterial infections made possible by the large disruptions in their epidemiology brought about by measures taken to reduce transmission of SARS CoV2 during the recent pandemic, including closure of schools.^11^ Taken together, these data emphasise the need to improve understanding of viral-bacterial interactions in the respiratory tract, if measures to prevent disease and optimise immunity are to be improved.

## Conclusions

Our study confirms that LAIV significantly increases Sp carriage density in previously LAIV-naive recipient young children and that LAIV transiently increases presumed household Sp transmission. Despite the relatively modest density increase, an increase in household Sp transmission events occurred, emphasizing the potential for respiratory viruses to transiently drive population level Sp carriage, likely through triggering Sp replication. Whole data mixed group analysis comparing vaccine exposed and unexposed periods produced more borderline results suggesting other covariates may also affect Sp transmission. These results highlight important knowledge gaps for understanding of Sp transmission that warrant further investigation and suggest that respiratory viral vaccines may play a beneficial role in reducing upper airway bacterial transmission and hence disease caused by these bacteria.

## Data Availability

All data produced in the present study are available upon reasonable request to the authors

## Acknowledgements

The authors thank the European Society for Paediatric Infectious Diseases, Bill & Melinda Gates Foundation and Pfizer for funding support. The authors also thank all study participants, paediatric research nurses, doctors and pharmacists at each study site, the Bristol Children’s Vaccine Centre laboratory and administration staff, Brandon Yeo (liquidbrain) and Jeremy Metz for their support in R-coding and Robin Marlow for continued support during this study. The views expressed in this publication are those of the authors.

## Supplemental Information

**Figure S1.**
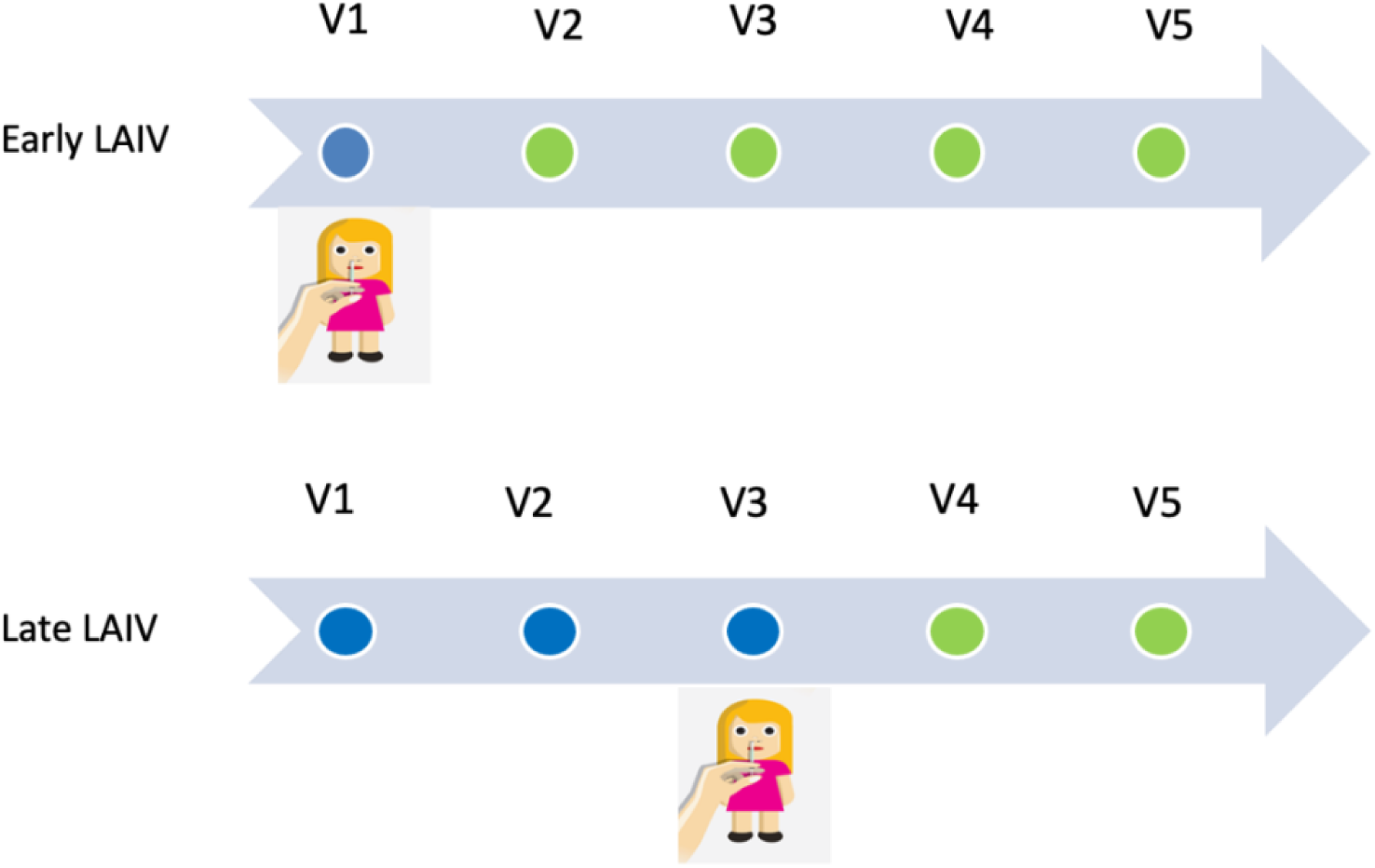
Study timeline for data and sample collection and LAIV administration. Baseline demographic data was collected at Visit 1 (V1). Clinical data and NP samples were collected from all participants at each study visit (V1-V5). In household families randomised into the early LAIV group, the index child received LAIV after samples were taken at visit 1. In household families randomised into the late LAIV group, the index child received LAIV after samples were taken at visit 3. The blue and green spots indicate study sampling times, blue, where index participants were LAIV-naïve and green where LAIV administration to index participants had taken place during previous visits.

**Figure S2:**
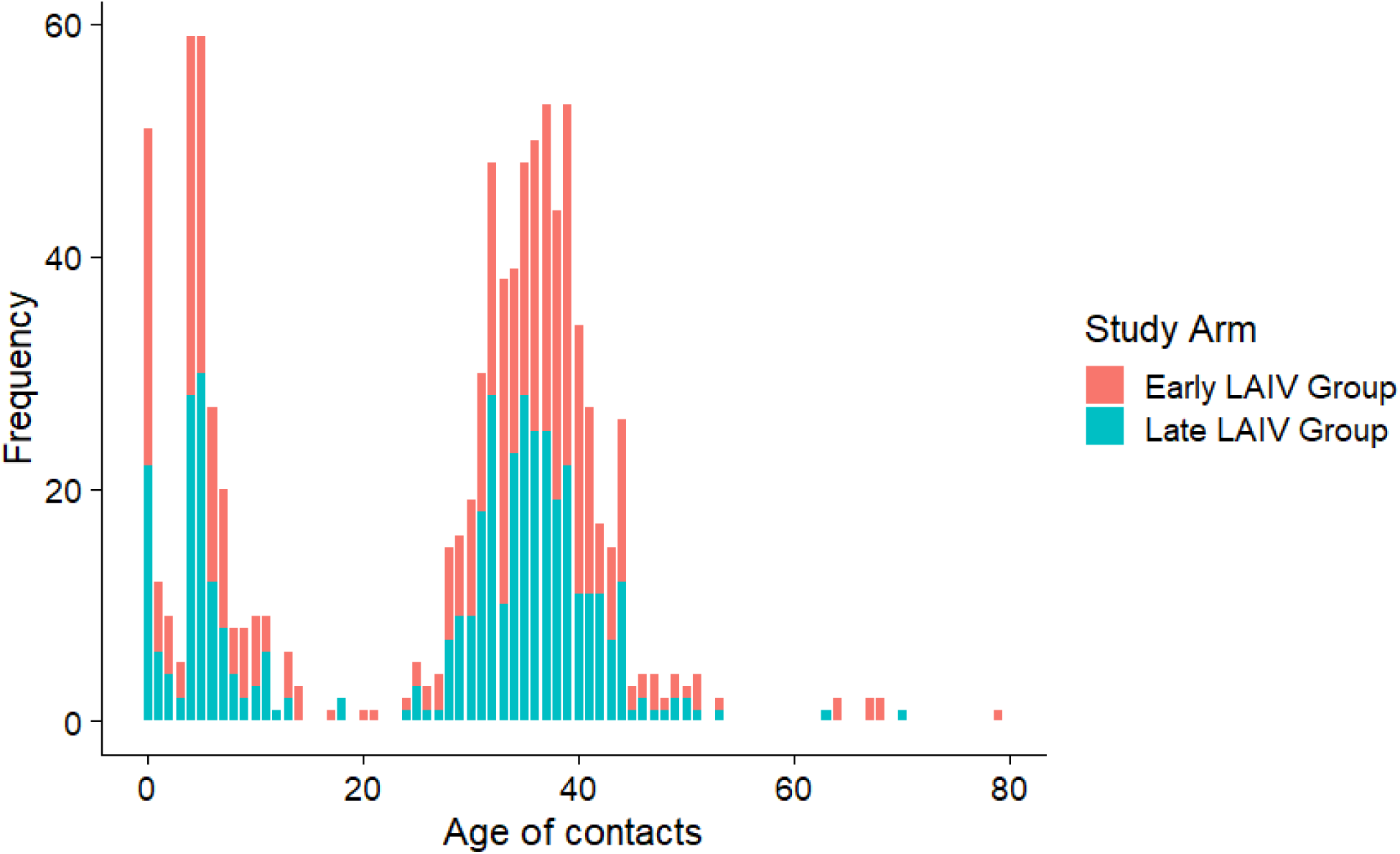
Age and frequency of the contacts of 2-year-old index children from the early (green) and late (red) LAIV study arms.

**Figure S3:**
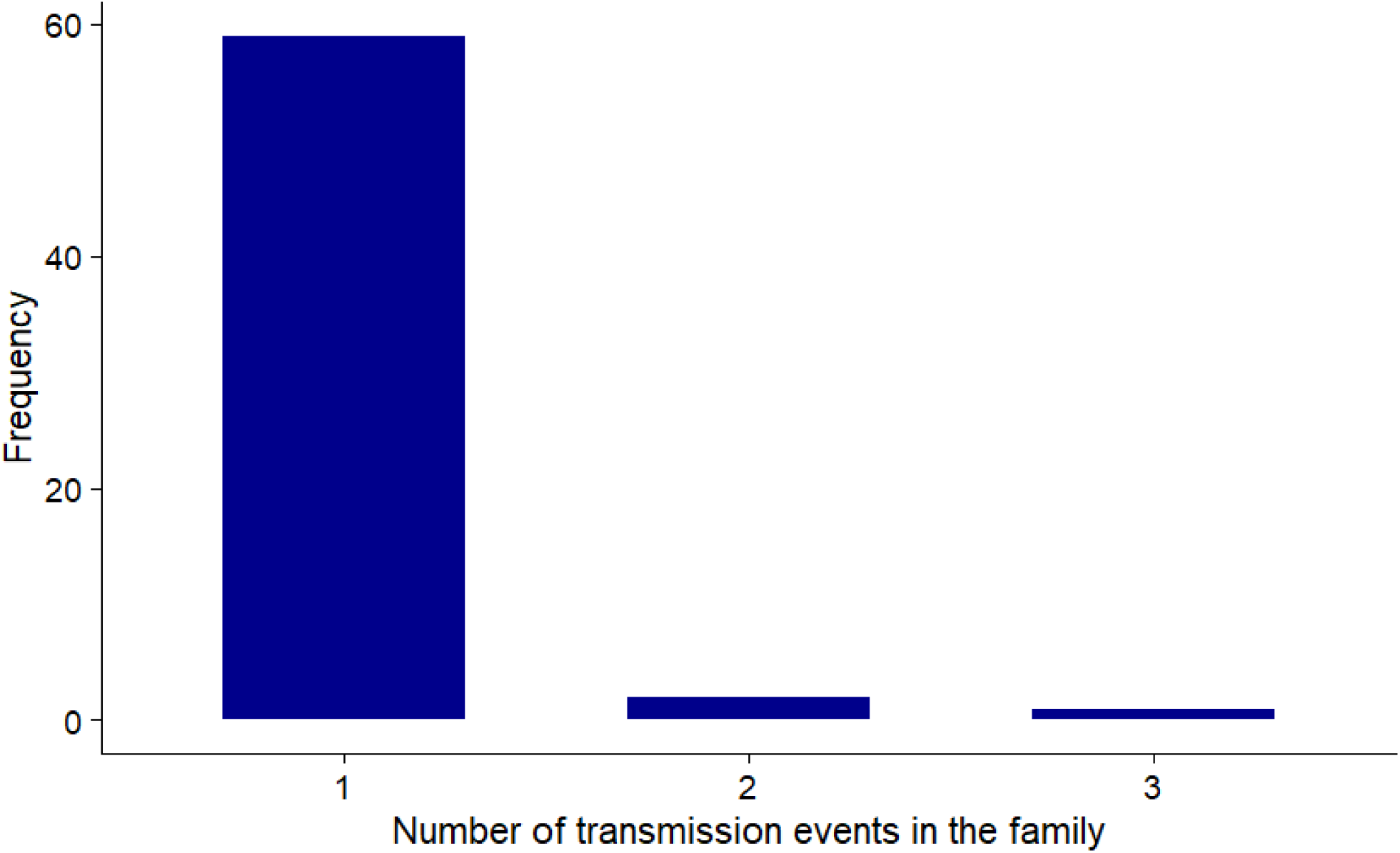
Histogram showing the number of pneumococcus transmission events among discordant pairs per family between visits 1 to 2

**Table S1:** Logistic regression mixed-effects model comparing change in pneumococcal density in 2-year-old index children at visit 2 (14 days) and visit 3 (28 days) between the early and late LAIV groups. We used a random intercept model to account for random effects between index children. In the early group, children received LAIV at visit 1 after baseline sampling, while in the late group, children did not receive LAIV until after sampling at visit 3.

| Visit number | Odds ratio | 95% CIs | p-value | S.D. of random effects | AIC of fixed effects model | AIC of mixed effects model |
| --- | --- | --- | --- | --- | --- | --- |
| Visit 1 -> 2 | 2.5 | 1.5 – 4.7 | <0.001 | <0.001 | 319 | 321 |
| Visit 1 -> 3 | 1.5 | 0.88 – 2.6 | 0.29 | <0.001 | 321 | 323 |

**Table S2:**
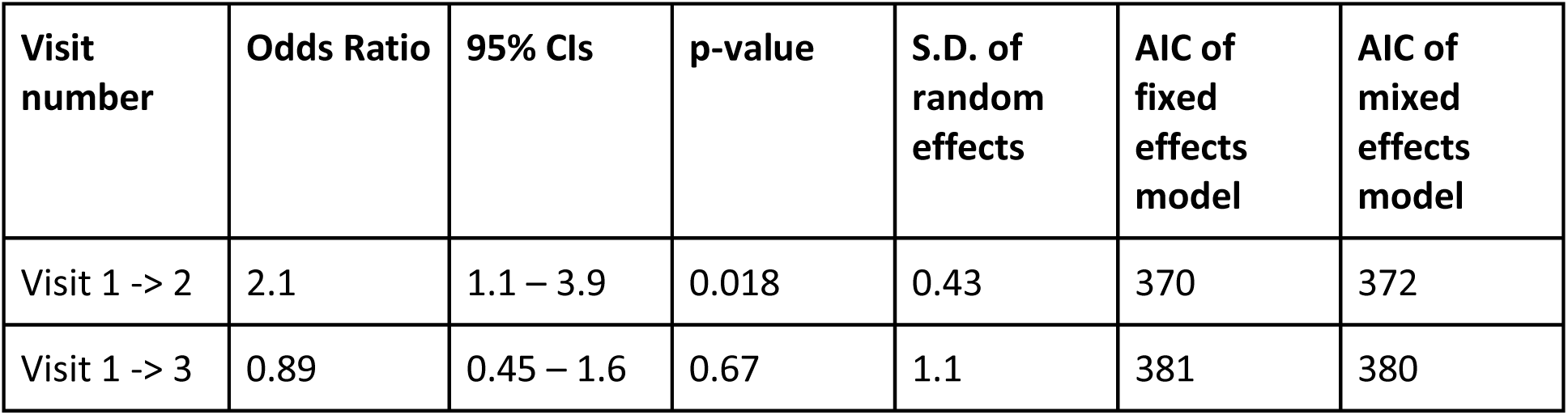
Logistic regression mixed-effects model comparing transmission events from 2-year-old index children carrying pneumococcus at baseline to family contacts at visit 2 (14 days) and visit 3 (28 days) between the early and late LAIV groups. We used a random intercept model to account for random effects between households participating in the study. In the early group, children received LAIV at visit 1 after baseline sampling, while in the late group, children did not receive LAIV until after sampling at visit 3.

| Visit number | Odds Ratio | 95% CIs | p-value | S.D. of random effects | AIC of fixed effects model | AIC of mixed effects model |
| --- | --- | --- | --- | --- | --- | --- |
| Visit 1 -> 2 | 2.1 | 1.1 – 3.9 | 0.018 | 0.43 | 370 | 372 |
| Visit 1 -> 3 | 0.89 | 0.45 – 1.6 | 0.67 | 1.1 | 381 | 380 |

**Table S3:**
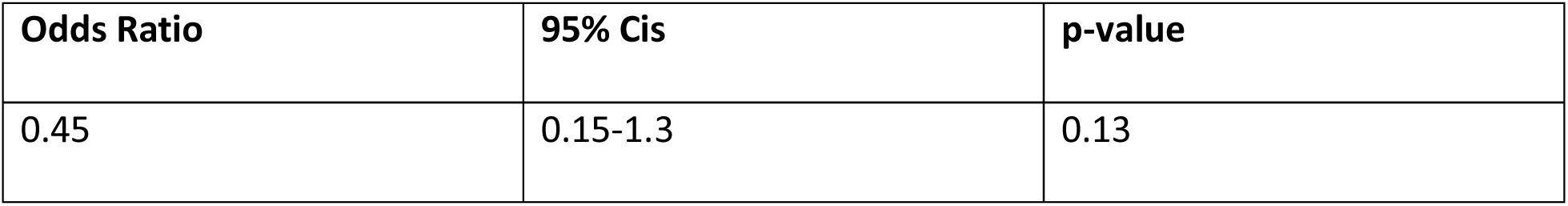
Logistic regression fixed-effects model comparing transmission events between early and late LAIV groups at visit 3 (28 days) from 2-year-old index children carrying pneumococcus at baseline to family contacts who were pneumococcus negative at visit 1 but positive at visits 2 and 3.

**Table S4:**
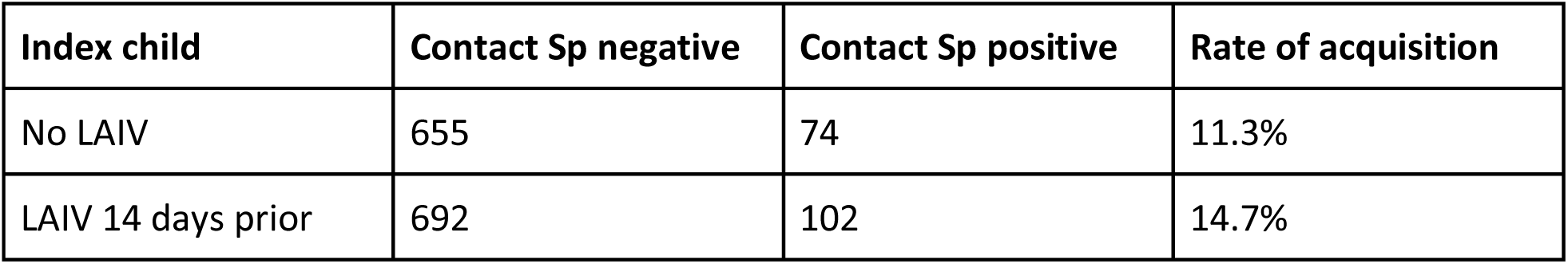
Comparison of rates of pneumococcal acquisition events occurring in families in which index 2-year-olds had not received LAIV (Control late LAIV group visits 1-2 and 2-3) with those in which these children had received LAIV 14 days previously (Early LAIV group visits 1-2 and Control late LAIV group visits 3-4). p = 0.06.

| Index child | Contact Sp negative | Contact Sp positive | Rate of acquisition |
| --- | --- | --- | --- |
| No LAIV | 655 | 74 | 11.3% |
| LAIV 14 days prior | 692 | 102 | 14.7% |

## Notes

### Competing Interest Statement

Brad Gessner is an employee of Pfizer holds stock in the company.

### Clinical Trial

ISRCTN10720581

### Funding Statement

This study was funded by the Bill & Melinda Gates Foundation and Pfizer.
I received a fellowship from the European Society of Paediatric Infectious Diseases

### Author Declarations

Ethical and governance approvals were granted by the UK Research Ethics Committee (Cambridgeshire and Hertfordshire) (17/EE/0351) and Health Research Authority

### Summary of Updates

Dear Sir or Madam, I am writing to enquire whether it would be possible to revise 2 tables and 2 figures (tables 2 and 3 and figures 4 and 5), and corresponding text within this paper. This comes after finding a bug within our code whilst conducting a separate analysis and realising results for a few participants (6) were not included when they should have been. In this corrected version I have also deleted an excess copy of S1 figure. With many thanks, Yours Faithfully, Dr Jane Metz

